# Vitamin B12 and Risk of Diabetes: New insight from Cross-Sectional and Longitudinal Analyses of the China Stroke Primary Prevention Trial (CSPPT)

**DOI:** 10.1101/2020.03.26.20044347

**Authors:** Lishun Liu, Xiao Huang, Binyan Wang, Yun Song, Tengfei Lin, Ziyi Zhou, Zhuo Wang, Yaping Wei, Huiyuan Guo, Ping Chen, Yan Yang, Wenhua Ling, Youbao Li, Xianhui Qin, Genfu Tang, Chengzhang Liu, Jianping Li, Yan Zhang, Pierre Zalloua, Xiaobin Wang, Yong Huo, Hao Zhang, Xiping Xu

## Abstract

**Introduction:** Previous studies in mostly Western populations, have yielded conflicting findings on the association of vitamin B12 with diabetes risk, in part, due to differences in study design and population characteristics. This study sought to examine the vitamin B12 – diabetes association in Chinese hypertensive adults by both cross-sectional and longitudinal analyses.

**Research Design and Methods:** This report included a total of 16699 participants from the China Stroke Primary Prevention Trial (CSPPT), with pertinent baseline and follow-up data. Diabetes mellitus (DM) was defined as either physician-diagnosed diabetes, the use of glucose-lowering drugs, or fasting blood glucose (FBG) ≥7.0 mmol/L. New-onset diabetes was defined as any new case of onset diabetes during the follow-up period or fasting blood glucose (FBG) ≥7.0 mmol/L at the exit visit.

**Results:** At baseline, there were 1872 (11.2%) diabetic patients; less than 1.5% had clinical B12 deficiency (<148.0 pmol/L). Over a median follow-up period of 4.5 years, there were 1589 (10.7%) cases of new-onset diabetes. Cross-sectional analyses showed a positive association between baseline vitamin B12 levels and FBG levels (β=0.18, 95%CI 0.15-0.21) and diabetes (OR=1.42, 95%CI 1.33-1.51). However, longitudinal analyses showed no association between baseline vitamin B12 and new-onset diabetes or changes in FBG levels. Among a subset of the sample (N=4366) with both baseline and exit B12 measurements, we found a positive association between an increase in B12 and an increase in FBG.

**Conclusions:** In this large Chinese hypertensive population mostly sufficient with vitamin B12, parallel cross-sectional and longitudinal analyses provided new insight into the conflicting findings of previous studies, and these results underscore the need for future studies to consider both baseline vitamin B12 and its longitudinal trajectory in order to better elucidate the role of vitamin B12 in the development of diabetes. Such findings, would have important clinical and public health implications.

## Introduction

Diabetes mellitus (DM) is a chronic metabolic disorder that has reached epidemic levels around the world ^1^. In China, there has been a sharp increase in diabetes prevalence in the past few decades and currently 11.4 million people have diabetes ^2^. From both clinical and public health perspectives, there is a critical need to develop cost-effective strategies to prevent diabetes. Vitamin B12 is a coenzyme in the one-carbon metabolic pathway, involved in the synthesis of methionine and pyrimidine and purine bases. Deficiencies in B12 and associated DNA damage and subsequent faulty repair are known to contribute to the development of vascular diseases, cancer, and some birth defects, and can lead to hyperhomocysteinemia. Often related to folic acid deficiency, vitamin B12 has been identified as a risk factor for both hypertension and atherosclerosis ^3^.

To date, most studies on B12 and DM have been centered on B12 deficiency among existing diabetic patients. The association between metformin use and low vitamin B12 levels has been supported by various levels of evidence^4^, but the risks and benefits of B12 on future risk of DM are not clear due to the inconsistent results of previous studies. Several observational studies have shown a link between low serum vitamin B12 concentrations and high body mass index (BMI), insulin resistance ^5^, and future type 2 diabetes mellitus (T2DM) onset. However, a cross-sectional study in a South Indian population showed that higher vitamin B12 levels decreased the risk of DM ^6^. Another longitudinal randomized control trial study showed no difference in the incidence of T2DM between the B12 supplemented group as compared to the non-supplemented control group^7^. The current study addresses an important yet controversial topic of whether vitamin B12 is associated with DM.

This current study was motivated by the findings of the U.S. National Health and Nutrition Examination Survey (NHANES)^8^ which showed that vitamin B12 levels in DM patients without metformin were significantly higher than those in the general population. However, the NHANES is a cross-sectional study, and in order to address whether B12 levels that are higher than the optimal range are a risk factor for developing DM, a prospective cohort study would be required to assess the temporal and dose-response relationship.

In this report, we analyzed a total of 16699 hypertensive participants of the China Stroke Primary Prevention Trial (CSPPT), with pertinent baseline data and a mean follow-up of 4.5 years. We performed both cross-sectional and longitudinal analyses with the aim of determining whether the findings of the NHANES could be replicated in a Chinese population, and furthermore, whether there is a prospective and dose-response association between baseline vitamin B12 levels and risk of new onset DM. Among a subset of the sample (N=4366) with both baseline and exit B12 measurements, we further analyzed the relationship between the change in B12 levels and the change in FBG levels from baseline to the exit visit.

## Research Design and Methods

### Participants and trial design

The parent study (the CSPPT) was approved by the Ethics Committee of the Institute of Biomedicine, Anhui Medical University, Hefei, China (Federal-wide Assurance Number: FWA00001263). All participants provided written, informed consent. A total of 20702 eligible participants, stratified by the methylenetetrahydrofolate reductase (*MTHFR*) C677T genotypes (CC, CT, or TT), were randomly assigned, in a 1:1 ratio, to one of two treatment groups: a daily oral dose of one tablet containing 10mg enalapril and 0.8mg folic acid (the enalapril-folic acid group), or a daily oral dose of one tablet containing 10mg enalapril only (the enalapril group). Participants were engaged in follow-up visits every three months.

A detailed description and the primary results of the CSPPT have been reported elsewhere^9-11^. Briefly, the CSPPT was a multi-community, randomized, double-blind, controlled trial conducted between May 19, 2008 and August 24, 2013 in 32 communities in China. Eligible participants were men and women aged 45-75 years with hypertension, defined as seated, resting systolic blood pressure (SBP)≥140 mmHg or diastolic blood pressure (DBP)≥90 mmHg at both the screening and recruitment visit or, who were taking antihypertensive medication. The major exclusion criteria included history of physician-diagnosed stroke, myocardial infarction (MI), heart failure, post-coronary revascularization, or congenital heart disease.

This report included 16699 hypertensive men and women from the CSPPT with baseline vitamin B12 data and pertinent baseline and follow-up data on diabetes status and covariables. As illustrated in the flow chart (Supplemental Fiigure 1), the final analyses excluded participants with missing values for baseline vitamin B12, baseline fasting blood glucose (FBG), exit FBG and with any missing data on the follow-up questionnaire. We also randomly selected a subset of the population (N=4366) to detect changes in B12 at the exit visit (Supplemental Table 1).

### Laboratory assays

Plasma vitamin B12 at baseline and folate at baseline and the exit visit were measured by a commercial lab using a chemiluminescent immunoassay (New Industrial, Shenzhen, China). Total homocysteine (tHcy), fasting lipids and FBG at baseline and the exit visit were measured using automatic clinical analyzers (Beckman Coulter, CA, USA) at the core lab of the National Clinical Research Center for Kidney Disease (Nanfang Hospital, Guangzhou, China).

### Outcomes

Patients were classified as diabetic if they self-reported a physician-diagnosis, or were using glucose-lowering medication, or when their FBG ≥7.0 mmol/L at baseline. New-onset diabetes was defined as a self-reported physician-diagnosis, or the use of glucose-lowering drugs during the follow-up period; or when FBG changed from <7.0 mmol/L at baseline to ≥7.0 mmol/L at the last study (exit) visit.

### Covariables

Covariables included known or suspected factors associated with vitamin B12 and/or DM based on existing literature including our own studies in the CSPPT, specifically, age, sex, *MTHFR* gene C677T polymorphisms, SBP and DBP at baseline, mean SBP and DBP during the treatment period, body mass index (BMI), study center, serum concentrations of folate, total homocysteine (tHcy), total cholesterol (TC), triglycerides (TG), high-density lipoprotein cholesterol (HDL-C), smoking status, alcohol consumption status, and self-reported meat consumption. Information on meat consumption was self-reported at baseline using a simple abbreviated semi-quantitative food frequency questionnaire. Participants were asked to report how often, on average, they eat meat every week. Possible response categories included “never”, “1-2 times/week”, “3-5 times/week” and “everyday”. B12 deficiency was defined as B12 <148.0pmol/L ^12^.

### Statistical analysis

Descriptive data are presented as the mean (SD) or proportions, as appropriate, for population characteristics according to baseline vitamin B12 quartiles. The significance of differences in population characteristics between groups was computed using two-sample t-tests, signed-rank tests, or Chi-squared tests, for continuous and categorical variables.

Logistic regression models were used to estimate the odds ratios (OR) and their 95% confidence intervals (CIs) of diabetes, given the exact onset of diabetes was not known and many new-onset DM cases were detected by fasting glucose levels at the exit visit. All analyses were conducted with adjustments for covariables. Finally, subgroup analyses were performed to evaluate possible effect modifications by the covariables on the association between B12 and DM including: sex (Male vs Female), age (<60 vs ≥60 years), *MTHFR* C677T polymorphism (CC vs CT vs TT), SBP (<160.0 vs ≥160.0 mmHg), DBP (<90 vs ≥90 mmHg), mean SBP during the treatment period (<140.0 vs ≥140.0 mmHg), mean DBP during the treatment period (<90 vs ≥90 mmHg), BMI (<25 vs ≥25 kg/m^2^), study center (Anqing vs Lianyungang), folate (<8 vs ≥8 ng/mL), tHcy (<15 vs ≥15 μmol/L), TC (<5.18 vs ≥5.18 mmol/L), TG (<1.7 vs ≥1.7 mmol/L), HDL-C (<1.16 vs ≥1.16 mmol/L) and treatment group (enalapril vs enalapril + folic acid). A two-tailed P<0.05 was considered significant in all analyses. All statistical analyses were performed using R software, version 3.6.0 (http://www.R-project.org/, accessed 26 April 2019).

## Results

### Study participants and baseline characteristics

Study participants had an average age of 60.0 years (SD 7.4), 6713 were male (40.2%) and 9986 were female (59.8%). Participants had an average B12 level of 295.9pmol/L (SD 91.5), 257(1.5%) participants had B12 deficiency, while 10468 (63.3%) participants did not consume meat. Mean FBG level was 5.8 mmol/L (SD 1.7) at baseline, and exit mean FBG level was 6.3 mmol/L (SD 2.0). At baseline, 1872 (11.2%) participants had DM and at the exit visit, there were 1589 (10.7%) cases of new-onset DM. When stratified by baseline vitamin B12 quartiles, FBG levels were the highest (6.0 mmol/L [SD 2.1]) in the fourth quartile (Q4). Table 1 shows that the average age and BMI of the participants in Q4 were lower than those of the other groups, but total cholesterol and triglyceride levels were higher than the other quartiles.

**Table 1.**
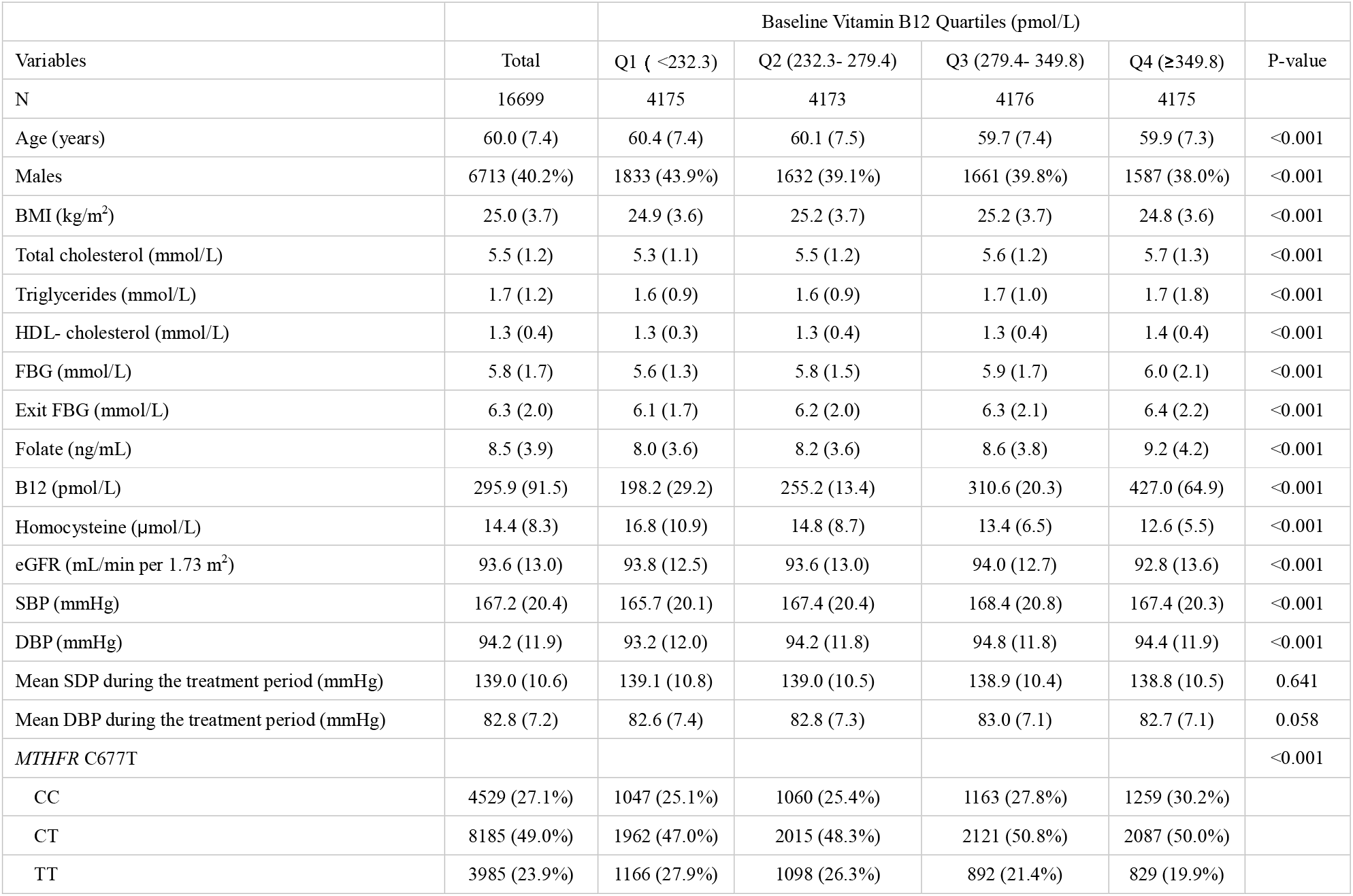
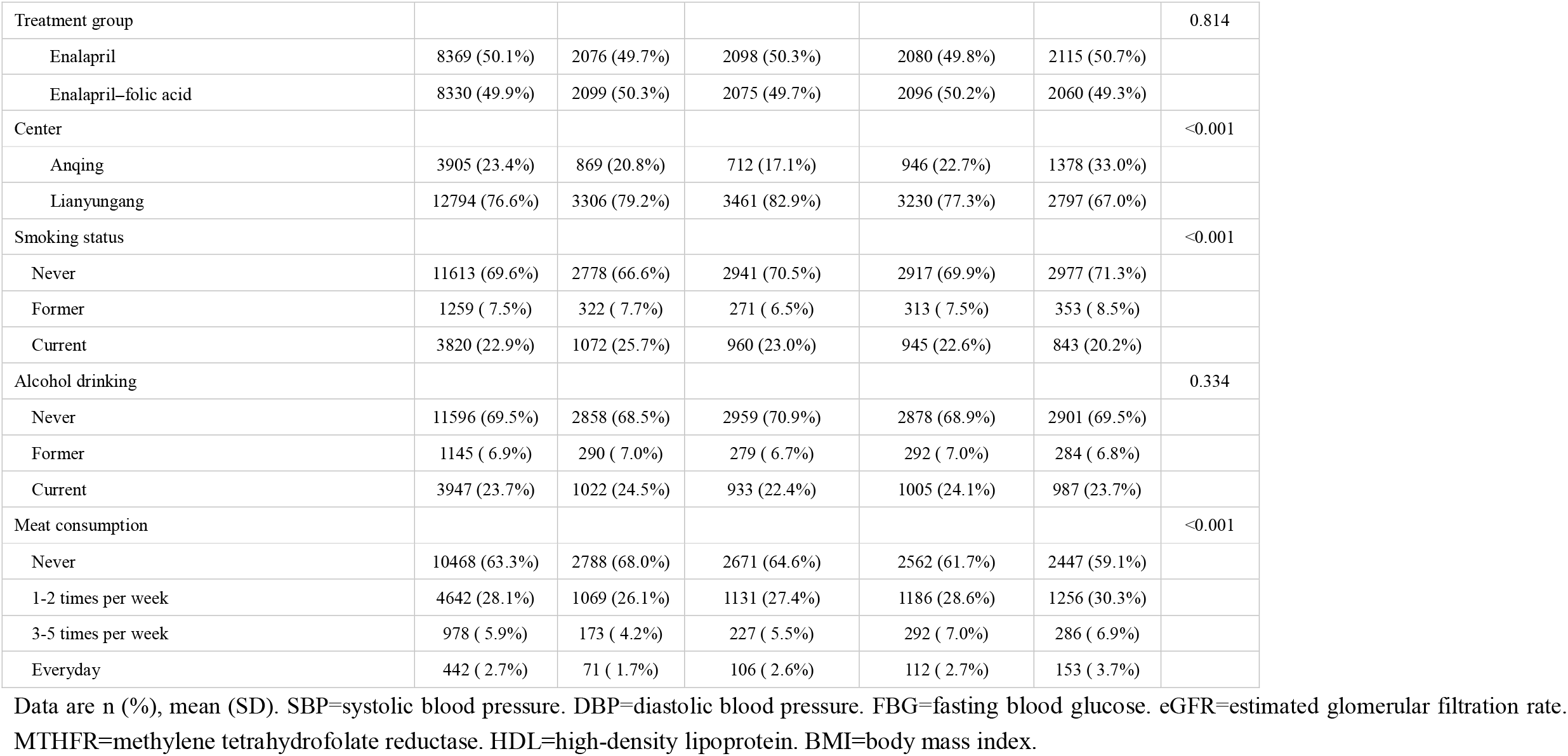
Baseline and follow-up characteristics of the study participants by baseline B12 quartiles.

### Cross-sectional analysis on baseline B12 and DM

From the cross-sectional analysis, B12 was found to be positively associated with DM (OR=1.42, 95%CI [1.33, 1.51], p<0.001) at baseline (Table 2). After stratifying by B12 quartiles, participants in Q4 were found to have the highest risk (OR=1.91, 95%CI [1.64, 2.23], p<0.001). Also, there was a positive association between B12 and FBG (β=0.18, 95%CI [0.15, 0.21], p<0.001) (Table 3). After stratifying by relevant covariables, we discovered an interaction between total cholesterol levels and vitamin B12 with DM (Supplemental Figure 2). For participants with total cholesterol levels ≥ 5.18 mmol/L and those with triglyceride levels ≥ 1.7 mmol/L, and also in females, B12 was more significantly associated with FBG levels (Supplemental Figure 3). No interaction was found between B12 and plasma folic acid or tHcy.

**Table 2.** Cross-sectional and longitudinal association between baseline vitamin B12 levels, and diabetes (DM) and new onset DM.

|  | Cross-sectional* |  |  |  |  | Longitudinal† |  |  |  |
| --- | --- | --- | --- | --- | --- | --- | --- | --- | --- |
|  | DM |  |  |  |  | new onset DM |  |  |  |
|  | N | Cases(%) | Non-adjusted | Adjusted |  | N | Cases(%) | Non-adjusted | Adjusted |
| Baseline B12 |  |  | OR (95%CI) p-value | OR (95%CI) p-value |  |  |  | OR (95%CI) p-value | OR (95%CI) p-value |
| Continuous (per IQR) | 16699 | 1872 (11.2) | 1.36 (1.28, 1.44) <0.001 | 1.42 (1.33, 1.51) <0.001 |  | 14827 | 1589 (10.7) | 1.02 (0.95, 1.09) 0.572 | 1.01 (0.94, 1.09) 0.734 |
| Quartiles |  |  |  |  |  |  |  |  |  |
| Q1 | 4175 | 353 ( 8.5) | 1 | 1 |  | 3707 | 391 (10.5) | 1 | 1 |
| Q2 | 4173 | 404 ( 9.7) | 1.15 (0.99, 1.34) 0.071 | 1.06 (0.90, 1.24) 0.482 |  | 3706 | 383 (10.3) | 0.99 (0.85, 1.14) 0.849 | 0.97 (0.83, 1.13) 0.726 |
| Q3 | 4176 | 493 (11.8) | 1.47 (1.27, 1.69) <0.001 | 1.34 (1.15, 1.56) <0.001 |  | 3707 | 417 (11.2) | 1.07 (0.92, 1.24) 0.387 | 1.05 (0.90, 1.23) 0.502 |
| Q4 | 4175 | 622 (14.9) | 1.83 (1.59, 2.11) <0.001 | 1.91 (1.64, 2.23) <0.001 |  | 3707 | 398 (10.7) | 1.02 (0.88, 1.18) 0.820 | 1.00 (0.86, 1.17) 0.982 |
| p for trend |  |  | <0.001 | <0.001 |  |  |  | 0.582 | 0.734 |

\*Adjusted for age, sex, methylene tetrahydrofolate reductase (MTHFR) gene C677T polymorphisms, systolic blood pressure (SBP) and diastolic blood pressure (DBP) at baseline, body mass index, study center, serum concentrations of folate, homocysteine, total cholesterol, triglycerides, and high-density lipoprotein cholesterol, smoking status, and alcohol consumption status.
† Adjusted for age, sex, methylene tetrahydrofolate reductase (MTHFR) gene C677T polymorphisms, systolic blood pressure (SBP) and diastolic blood pressure (DBP) at baseline, mean SBP and DBP during the treatment period, body mass index, study center, baseline serum concentrations of folate, homocysteine, fasting blood glucose (FBG), total cholesterol, triglycerides, and high-density lipoprotein cholesterol, treatment group, smoking status, and alcohol consumption status.
OR=odds ratio,

**Table 3.** Cross-sectional and longitudinal association between baseline vitamin B12 and baseline FBG, exit FBG and change in FBG (^△^FBG).

|  | Cross-sectional* |  |  | Longitudinal† |  |  |  |  |  |
| --- | --- | --- | --- | --- | --- | --- | --- | --- | --- |
| | | Baseline FBG | | | Exit FBG | | | ΔFBG | |
|  | Mean(SD) | Non-adjusted | Adjusted | Mean(SD) | Non-adjusted | Adjusted | Mean(SD) | Non-adjusted | Adjusted |
| **Baseline B12** | | $\beta$ (95%CI) p-value | $\beta$ (95%CI) p-value | | $\beta$ (95%CI) p-value | $\beta$ (95%CI) p-value | | $\beta$ (95%CI) p-value | $\beta$ (95%CI) p-value |
| **Continuous** | 5.8(1.7) | 0.16 (0.13, 0.19) | 0.18 (0.15, 0.21) | 6.3 (2.0) | 0.13 (0.09, 0.17) | 0.02 (-0.01, 0.06) | 0.5 (1.6) | -0.02 (-0.05, 0.01) | -0.02 (-0.05, 0.01) |
| **(per IQR)** |  | <0.001 | <0.001 |  | <0.001 | 0.173 |  | 0.163 | 0.154 |
| **Quartiles** |  |  |  |  |  |  |  |  |  |
| **Q1** | 5.6(1.3) | 0 | 0 | 6.1 (1.7) | 0 | 0 | 0.5 (1.5) | 0 | 0 |
| **Q2** | 5.8(1.5) | 0.11 (0.04, 0.18) | 0.04 (-0.03, 0.11) | 6.2 (2.0) | 0.07 (-0.01, 0.16) | 0.01 (-0.06, 0.08) | 0.5 (1.7) | -0.04 (-0.11, 0.03) | -0.01 (-0.08, 0.06) |
|  |  | 0.002 | 0.239 |  | 0.094 | 0.774 |  | 0.241 | 0.729 |
| **Q3** | 5.9(1.7) | 0.23 (0.16, 0.31) | 0.18 (0.11, 0.24) | 6.3 (2.1) | 0.19 (0.10, 0.28) | 0.04 (-0.03, 0.11) | 0.5 (1.7) | -0.05 (-0.12, 0.02) | -0.02 (-0.09, 0.05) |
|  |  | <0.001 | <0.001 |  | <0.001 | 0.293 |  | 0.144 | 0.523 |
| **Q4** | 5.9(2.1) | 0.31 (0.24, 0.38) | 0.34 (0.27, 0.41) | 6.4 (2.2) | 0.23 (0.14, 0.32) | 0.02 (-0.05, 0.10) | 0.4 (1.7) | -0.05 (-0.12, 0.02) | -0.06 (-0.13, 0.01) |
|  |  | <0.001 | <0.001 |  | <0.001 | 0.568 |  | 0.168 | 0.109 |
| **p for trend** |  | <0.001 | <0.001 |  | <0.001 | 0.437 |  | 0.162 | 0.109 |

\*Adjusted for age, sex, methylene tetrahydrofolate reductase (MTHFR) gene C677T polymorphisms, systolic blood pressure (SBP) and diastolic blood pressure (DBP) at baseline, body mass index, study center, serum concentrations of folate, homocysteine, total cholesterol, triglycerides, and high-density lipoprotein cholesterol, smoking status, and alcohol consumption status.
†Adjusted for age, sex, methylene tetrahydrofolate reductase (MTHFR) gene C677T polymorphisms, systolic blood pressure (SBP) and diastolic blood pressure (DBP) at baseline, mean SBP and DBP during the treatment period, body mass index, study center, baseline serum concentrations of folate, homocysteine, fasting blood glucose (FBG), total cholesterol, triglycerides, and high-density lipoprotein cholesterol, treatment group, smoking status, and alcohol consumption status.

### Longitudinal analyses on baseline B12 and new onset DM

Longitudinal analyses did not show an association between baseline B12 and new onset DM (OR=1.01, 95%CI [0.94, 1.09], p=0.734) (Table 2), change in FBG (β= -0.02, 95%CI [-0.05, 0.01], p=0.154), or exit FBG (β=0.02, 95%CI[-0.01, 0.06], p=0.173) after making additional adjustments for baseline FBG. (Table 3). After stratifying by relevant covariables, no interaction was found between vitamin B12 with new onset DM, exit FBG or change in FBG (Supplemental Figure 2, Supplemental Figure 3)

### Longitudinal analyses on change in B12 levels and change in FBG levels

Among a subset of the sample (N=4366) with both baseline and exit B12 measurements, we further analyzed the relationship between change in B12 levels and change in FBG levels from the baseline to the exit visit. We found a dose-response relationship between change in B12 and change in FBG levels (Table 4, Figure 1).

**Table 4.** Longitudinal analyses on change in vitamin B12 (^△^B12) and change in FBG (^△^FBG) from baseline to exit visit*.

| Exposure | | | ΔFBG (μmol/L) | | | |
| --- | --- | --- | --- | --- | --- | --- |
|  |  |  | Non-adjusted model |  | Adjusted model† |  |
| ΔB12 | N | Mean (SD) | $\beta$ ( 95%CI ) | p-value | $\beta$ ( 95%CI ) | p-value |
| Continuous (per IQR) | 4366 | -15.4(299.5) | 0.02(0.00,0.04) | 0.018 | 0.01(0.00,0.03) | 0.127 |
| Quartiles |  |  |  |  |  |  |
| Q1 | 1092 | -149.1(155.3) | 1 |  | 1 |  |
| Q2 | 1091 | -38.4(13.5) | 0.05(-0.09,0.19) | 0.479 | -0.01(-0.13,0.12) | 0.920 |
| Q3 | 1091 | 6.5(13.5) | 0.23(0.09,0.37) | 0.001 | 0.11(-0.02,0.23) | 0.096 |
| Q4 | 1092 | 119.2(545.2) | 0.34(0.20,0.48) | <0.001 | 0.21(0.08,0.33) | 0.001 |
| p for trend |  |  |  | <0.001 |  | <0.001 |

\* This subsample included a total of 4366 subjects with both baseline and exit FBG and B12 measurements.
†Adjusted for age, sex, methylene tetrahydrofolate reductase (MTHFR) gene C677T polymorphisms, systolic blood pressure (SBP) and diastolic blood pressure (DBP) at baseline, mean SBP and DBP during the treatment period, body mass index, study center, baseline serum concentrations of folate, homocysteine, fasting blood glucose (FBG), B12, total cholesterol, triglycerides, and high-density lipoprotein cholesterol, treatment group, smoking status, and alcohol consumption status.

**Figure 1.**
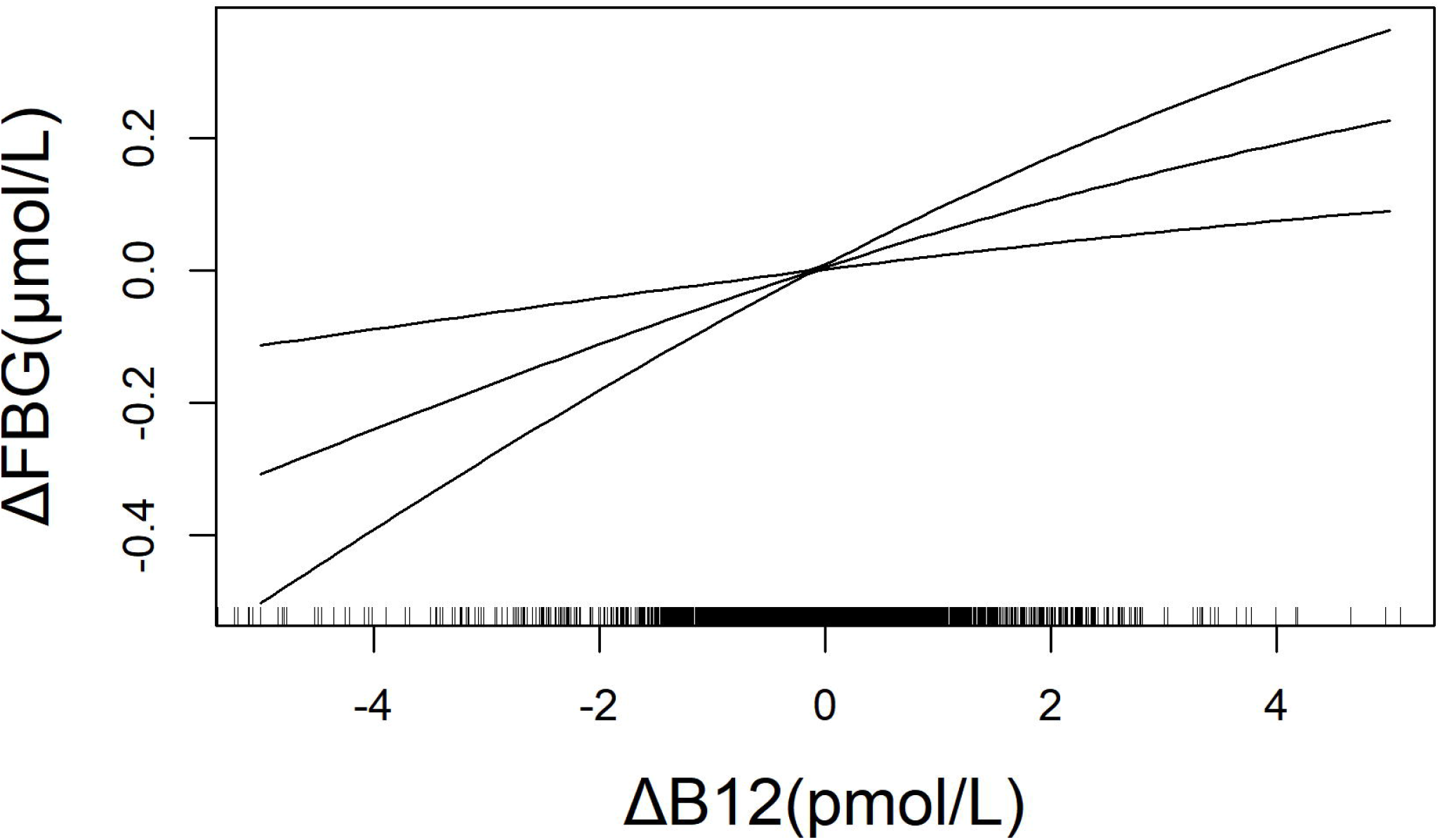
Multivariable-adjusted Smoothing curves of change in B12 and change in FBG in subsample included a total of 4366 subjects with both baseline and exit FBG and B12 measurements*. *Adjusted for age, sex, methylene tetrahydrofolate reductase (MTHFR) gene C677T polymorphisms, systolic blood pressure (SBP) and diastolic blood pressure (DBP) at baseline, mean SBP and DBP during the treatment period, body mass index, study center, baseline serum concentrations of folate, homocysteine, fasting blood glucose (FBG), B12, total cholesterol, triglycerides, and high-density lipoprotein cholesterol, treatment group, smoking status, and alcohol consumption status.

## Discussion

This is the first time that the relationship between vitamin B12 and DM has been explored in a Chinese hypertensive population via both cross-sectional and longitudinal analyses. We confirmed the findings of the NHANES, showing a cross-sectional positive association between B12 with DM at baseline in this Chinese population. Furthermore, our longitudinal analyses demonstrated that there was no association between baseline vitamin B12 levels and new onset DM risk. Our study has contributed new insights on the B12 and DM association and has helped to explain inconsistent findings in previous studies.

### First, the B12 and DM association depends on the population characteristics

Most previous studies on the association of B12 and DM, were centered on B12 deficiency among existing DM patients with the use of metformin. The association between metformin use and low vitamin B12 levels has been supported by various levels of evidence ^4^. Most of those studies were conducted in older populations, where B12 deficiency is more likely ^8^. In contrast, our study was conducted in a Chinese hypertensive population who were relatively young (45-75 years at baseline), mostly free from DM at baseline, and mostly B12 sufficient. Only 6.9% of the study participants with DM reported using metformin.

### Second, the B12 and DM association depends on the study design and type of analyses

From the cross-sectional analyses: Moen, G. H. et al. found that vitamin B12 may have a causal effect on fasting glucose^13^. However, Jayashri, R. et al. found that the levels of vitamin B12 decreased with increasing severity of glucose tolerance^6^. Margalit, I. et al. found no significant difference in blood sugar between the B12 deficient group and the non-deficient group^14^. From the longitudinal analyses and the randomized trials: Looker, H.C. et al. found that vitamin B12 was positively associated with all-cause mortality, and death from diabetes/nephropathy^15^. Song, Y. et al. found that daily supplementation with folic acid and vitamins B6 and B12 did not reduce the risk of developing type 2 diabetes among women at high risk for CVD^7^. Kwok, T. et al. found that vitamin B12 supplementation did not prevent cognitive decline in older diabetic patients with borderline vitamin B12 status^16^. From a systematic review: Rafnsson, S. B. et al. found that current data do not support vitamin B12 supplementation to reduce the risk of cardiovascular diseases or diabetes^17^.

Our study was the first to perform and report findings from both cross-sectional and longitudinal analyses in the same population. In the cross-sectional analysis, we found an independent, positive association between baseline B12 levels and DM and FBG. These results persisted even after we adjusted for relevant covariables. This finding is consistent with the NHANES study^8^. In the longitudinal analyses, we did not find any association between baseline B12 and new onset DM risk. This finding is consistent with the Women’s Antioxidant and Folic Acid Cardiovascular Study, where women aged ≥40 years with a history of cardiovascular disease, who were free of DM at baseline, were supplemented with either a combination pill consisting of folic acid, pyridoxine and B12, or a placebo. After a median follow-up of 7.3 years, no difference in incident T2DM was found between the two groups^7^. Another longitudinal study^18^ in Japan also reported similar null results. Taken together, our longitudinal analyses and that of others did not support an association between B12 and new-onset DM. These findings underscore that cross-sectional associations need to be confirmed by prospective studies and clinical trials, because the former is more likely to be subject to many drawbacks, including reverse causality.

### Third, our findings have clinical implications

The role of B12 in DM varied by patient characteristics. Most previous studies have shown that B12 supplementation is necessary in elderly diabetic patients with low B12 levels or in diabetic patients with long-term metformin use ^8 19 20^. Our study, along with other longitudinal studies, however, does not support the routine use of vitamin B12 supplementation to reduce the risk of new onset DM^17^ in relatively young patients with no evidence of B12 deficiency. Moreover, a meta-analysis by Valdés-Ramos et al. indicated no recommendation for the use of vitamin supplements in patients with T2DM ^21^. Of note, the research of Helen C. Looker et al. ^15^ showed that vitamin B12 was positively associated with all-cause mortality and death from diabetes/nephropathy, and previous data also indicated that elevated serum vitamin B12 levels are a predictive factor for mortality in elderly patients with cancer^22^. Salles, N. et al.^23^ and Hemmersbach-Miller, M. et al.^24^ reported that higher vitamin B12 levels might also be a marker to assess a higher risk of mortality in elderly patients. Vitamin B12 can also accelerate decline in renal function and increase the risk of cardiovascular events in patients with impaired renal function ^25 26^. Zeitlin et al. ^27^ also suggest that for elderly people, vitamin B12 supplementation should not be routinely provided unless there are clear indications for doing so (a deficiency state), and then, to only replace enough B12 to correct the deficiency. Through our research and analysis, we found that B12 may still have a correlation with blood glucose or diabetes mellitus, and the disappearance of this correlation in the longitudinal analysis may be due to the relative changes in the observation age, the decrease of B12 levels and the increase of FBG levels over time. Therefore, the relationship between the changes in indicators need to be observed to reflect their real results. We found that the change in B12 levels and the change in FBG levels showed a positive vitamin B12-FBG association in the subsample. In addition, we repeated the previous analysis with this subsample, and found the results were consistent with those of the previous analysis (Supplemental Table S2).

The present study had some limitations. First, this study focused on Chinese adults with hypertension, so the generalizability of the results to other populations remains to be determined. Second, new-onset DM was not a primary outcome or a prespecified outcome of the CSPPT. We did not obtain FBG measurements at the scheduled follow-up visits, nor did we measure HbA1c or perform glucose tolerance tests at baseline or during the follow-up visits. Therefore, it is possible that we have underestimated the incidence of new-onset DM in the CSPPT. Nevertheless, we believe that any potential underestimation of new-onset DM should be non-differential, and therefore should not significantly affect the results. Finally, we only measured B12 levels on a small subset of the population at the exit visit and were unable to examine B12 dynamics during the follow-up period of the CSPPT.

## Conclusion

Among a population of adults with hypertension in China without a history of stroke or MI who were mostly B12 sufficient, there was a dose-response association of vitamin B12 levels with the risk of DM based on cross-sectional analyses at baseline. There was no prospective relationship between baseline B12 and new onset DM in the longitudinal analyses. However, in a subsample, a positive vitamin B12-FBG association was shown by the change in B12 levels and the change in FBG levels. Our findings illustrate a clear discrepancy in results from the cross-sectional and longitudinal analyses even from the same study population, and underscore the need to consider both baseline and longitudinal changes between vitamin B12 and FBG in order to better elucidate the role of vitamin B12 in the development of diabetes. If further studies confirm such findings, this will have an important impact on clinical and public health.

## Data Availability

All data included in this study are available upon request by contact with the corresponding author.

## Acknowledgements

The China Stroke Primary Prevention Trial (CSPPT) was jointly supported by Shenzhen AUSA Pharmed (Shenzhen, China) and national, provincial and private funding, including from the Major State Basic Research Development Program of China (973 program; grant no. 2102 CB517703); the National Science and Technology Major Projects Specialized for “Innovation and Development of Major New Drugs” during the 12th Five-year Plan Period: the China Stroke Primary Prevention Trial (grant no. zx09101105), a Clinical Center grant (no. zx09401013); the Projects of the National Natural Science Foundation of China (grant no. 81473052, 81441091, and 81402735); the National Clinical Research Center for Kidney Disease, Nanfang Hospital, Nanfang Medical University, Guangzhou, China; the State Key Laboratory for Organ Failure Research, Nanfang Hospital; and research grants from the Department of Development and Reform, Shenzhen Municipal Government (grant no. SFG 20201744). The funding organizations and/or sponsor participated in the study design, but had no role in the conduct of the study; collection, management, analysis, and interpretation of the data; preparation, review, or approval of the manuscript; or the decision to submit the manuscript for publication.

## Contributors

Xiping Xu, Yong Huo and Hao Zhang critically revised the protocol for research design. Lishun Liu, Xiao Huang, Yun Song, Tengfei Lin, Ziyi Zhou, Zhuo Wang, Ping Chen, and Genfu Tang were responsible for implementation onsite. Lishun Liu, Xiao Huang and Chengzhang Liu performed the statistical analyses. Lishun Liu and Xiao Huang drafted the manuscript. Xiaobin Wang, Pierre Zalloua, Yan Yang, Wenhua Ling, Jianping Li, Yan Zhang, Youbao Li, Xianhui Qin and Binyan Wang developed the methodological approach. All authors contributed to the conception and design and approved the final version of the manuscript.

---

Supplemental Figure 1. Flow chart of participants for the study

Supplemental Figure 2. Forest plot on the cross-sectional and longitudinal associations between baseline B12, baseline DM and new onset DM.

*Adjusted for age, sex, methylene tetrahydrofolate reductase *(MTHFR)* gene C677T polymorphisms, systolic blood pressure (SBP) and diastolic blood pressure (DBP) at baseline, body mass index, study center, serum concentrations of folate, homocysteine, total cholesterol, triglycerides, and high-density lipoprotein cholesterol, smoking status, and alcohol consumption status.

† Adjusted for age, sex, methylene tetrahydrofolate reductase (MTHFR) gene C677T polymorphisms, systolic blood pressure (SBP) and diastolic blood pressure (DBP) at baseline, mean SBP and DBP during the treatment period, body mass index, study center, baseline serum concentrations of folate, homocysteine, fasting blood glucose (FBG), total cholesterol, triglycerides, and high-density lipoprotein cholesterol, treatment group, smoking status, and alcohol consumption status.

Supplemental Figure 3. Forest plot on the cross-sectional and longitudinal associations between baseline B12, baseline FBG, exit FBG and ^△^FBG

*Adjusted for age, sex, methylene tetrahydrofolate reductase *(MTHFR*) gene C677T polymorphisms, systolic blood pressure (SBP) and diastolic blood pressure (DBP) at baseline, mean SBP and DBP during the treatment period, body mass index, study center, serum concentrations of folate, homocysteine, total cholesterol, triglycerides, and high-density lipoprotein cholesterol, treatment group, smoking status, and alcohol consumption status.

† Adjusted for age, sex, methylene tetrahydrofolate reductase (MTHFR) gene C677T polymorphisms, systolic blood pressure (SBP) and diastolic blood pressure (DBP) at baseline, mean SBP and DBP during the treatment period, body mass index, study center, baseline serum concentrations of folate, homocysteine, fasting blood glucose (FBG), total cholesterol, triglycerides, and high-density lipoprotein cholesterol, treatment group, smoking status, and alcohol consumption status.

## References

1. Chatterjee S, Khunti K, Davies MJ. Type 2 diabetes. Lancet 2017;389(10085):2239–51. doi: 10.1016/S0140-6736(17)30058-2 [published Online First: 2017/02/10]

2. Chinese Diabetes S, National Offic for Primary Diabetes C. [National guidelines for the prevention and control of diabetes in primary care(2018)]. Zhonghua Nei Ke Za Zhi 2018;57(12):885–93. doi: 10.3760/cma.j.issn.0578-1426.2018.12.003 [published Online First: 2018/11/30]

3. O’Leary F, Samman S. Vitamin B12 in health and disease. Nutrients 2010;2(3):299–316. doi: 10.3390/nu2030299 [published Online First: 2010/03/05]

4. Yang W, Cai XL, Wu H, et al. Associations between metformin use and vitamin B12 level, anemia and neuropathy in patients with diabetes: a meta-analysis. J Diabetes 2019 doi: 10.1111/1753-0407.12900 [published Online First: 2019/01/07]

5. Knight BA, Shields BM, Brook A, et al. Lower Circulating B12 Is Associated with Higher Obesity and Insulin Resistance during Pregnancy in a Non-Diabetic White British Population. PLoS One 2015;10(8):e0135268. doi: 10.1371/journal.pone.0135268 [published Online First: 2015/08/20]

6. Jayashri R, Venkatesan U, Rohan M, et al. Prevalence of vitamin B12 deficiency in South Indians with different grades of glucose tolerance. Acta Diabetol 2018;55(12):1283–93. doi: 10.1007/s00592-018-1240-x [published Online First: 2018/10/15]

7. Song Y, Cook NR, Albert CM, et al. Effect of homocysteine-lowering treatment with folic Acid and B vitamins on risk of type 2 diabetes in women: a randomized, controlled trial. Diabetes 2009;58(8):1921–8. doi: 10.2337/db09-0087 [published Online First: 2009/06/02]

8. Reinstatler L, Qi YP, Williamson RS, et al. Association of biochemical B□□ deficiency with metformin therapy and vitamin B□□ supplements: the National Health and Nutrition Examination Survey, 1999-2006. Diabetes Care 2012;35(2):327–33. doi: 10.2337/dc11-1582 [published Online First: 2011/12/16]

9. Huo Y, Li J, Qin X, et al. Efficacy of folic acid therapy in primary prevention of stroke among adults with hypertension in China: the CSPPT randomized clinical trial. JAMA 2015;313(13):1325–35. doi: 10.1001/jama.2015.2274

10. Huang X, Li Y, Li P, et al. Association between percent decline in serum total homocysteine and risk of first stroke. Neurology 2017;89(20):2101–07. doi: 10.1212/WNL.0000000000004648 [published Online First: 2017/10/17]

11. Huang X, Qin X, Yang W, et al. MTHFR Gene and Serum Folate Interaction on Serum Homocysteine Lowering: Prospect for Precision Folic Acid Treatment. Arterioscler Thromb Vasc Biol 2018;38(3):679–85. doi: 10.1161/ATVBAHA.117.310211 [published Online First: 2018/01/27]

12. Hunt A, Harrington D, Robinson S. Vitamin B12 deficiency. BMJ 2014;349:g5226. doi: 10.1136/bmj.g5226 [published Online First: 2014/09/04]

13. Moen GH, Qvigstad E, Birkeland KI, et al. Are serum concentrations of vitamin B-12 causally related to cardiometabolic risk factors and disease? A Mendelian randomization study. Am J Clin Nutr 2018;108(2):398–404. doi: 10.1093/ajcn/nqy101

14. Margalit I, Cohen E, Goldberg E, et al. Vitamin B12 Deficiency and the Role of Gender: A Cross-Sectional Study of a Large Cohort. Ann Nutr Metab 2018;72(4):265–71. doi:10.1159/000488326 [published Online First: 2018/03/29]

15. Looker HC, Fagot-Campagna A, Gunter EW, et al. Homocysteine and vitamin B(12) concentrations and mortality rates in type 2 diabetes. Diabetes Metab Res Rev 2007;23(3):193–201. doi: 10.1002/dmrr.660

16. Kwok T, Lee J, Ma RC, et al. A randomized placebo controlled trial of vitamin B12 supplementation to prevent cognitive decline in older diabetic people with borderline low serum vitamin B12. Clin Nutr 2017;36(6):1509–15. doi: 10.1016/j.clnu.2016.10.018 [published Online First: 2016/11/09]

17. Rafnsson SB, Saravanan P, Bhopal RS, et al. Is a low blood level of vitamin B12 a cardiovascular and diabetes risk factor? A systematic review of cohort studies. Eur J Nutr 2011;50(2):97–106. doi: 10.1007/s00394-010-0119-6 [published Online First: 2010/06/29]

18. Eshak ES, Iso H, Muraki I, et al. Among the water-soluble vitamins, dietary intakes of vitamins C, B2 and folate are associated with the reduced risk of diabetes in Japanese women but not men. Br J Nutr 2019:1–22. doi: 10.1017/S000711451900062X [published Online First: 2019/03/21]

19. Gupta K, Jain A, Rohatgi A. An observational study of vitamin b12 levels and peripheral neuropathy profile in patients of diabetes mellitus on metformin therapy. Diabetes Metab Syndr 2018;12(1):51–58. doi: 10.1016/j.dsx.2017.08.014 [published Online First: 2017/08/25]

20. Kwok T, Lee J, Ma RC, et al. A randomized placebo controlled trial of vitamin B. Clin Nutr 2017;36(6):1509–15. doi: 10.1016/j.clnu.2016.10.018 [published Online First:2016/10/27]

21. Valdés-Ramos R, Guadarrama-López AL, Martínez-Carrillo BE, et al. Vitamins and type 2 diabetes mellitus. Endocr Metab Immune Disord Drug Targets 2015;15(1):54–63.

22. Geissbühler P, Mermillod B, Rapin CH. Elevated serum vitamin B12 levels associated with CRP as a predictive factor of mortality in palliative care cancer patients: a prospective study over five years. J Pain Symptom Manage 2000;20(2):93–103.

23. Salles N, Herrmann F, Sakbani K, et al. High vitamin B12 level: a strong predictor of mortality in elderly inpatients. J Am Geriatr Soc 2005;53(5):917–8. doi: 10.1111/j.1532-5415.2005.53278_7.x

24. Hemmersbach-Miller M, Conde-Martel A, Betancor-León P. Vitamin B as a predictor of mortality in elderly patients. J Am Geriatr Soc 2005;53(11):2035–6. doi: 10.1111/j.1532-5415.2005.00479_2.x

25. Spence JD, Yi Q, Hankey GJ. B vitamins in stroke prevention: time to reconsider. Lancet Neurol 2017;16(9):750–60. doi: 10.1016/S1474-4422(17)30180-1 [published Online First: 2017/08/18]

26. House AA, Eliasziw M, Cattran DC, et al. Effect of B-vitamin therapy on progression of diabetic nephropathy: a randomized controlled trial. JAMA 2010;303(16):1603–9. doi: 10.1001/jama.2010.490 [published Online First: 2010/04/29]

27. Zeitlin A, Frishman WH, Chang CJ. The association of vitamin b 12 and folate blood levels with mortality and cardiovascular morbidity incidence in the old old: the Bronx aging study. Am J Ther 1997;4(7-8):275-81. [published Online First: 1997/07/01]

